# Evaluating acute medical service performance against assessment time metrics: the Society for Acute Medicine Benchmarking Audit 2023 (SAMBA23)

**DOI:** 10.1101/2024.07.15.24310421

**Authors:** Catherine Atkin, Chris Subbe, Mark Holland, Ragit Varia, Tim Cooksley, Adnan Gebril, Adrian Kennedy, Thomas Knight, Daniel Lasserson

## Abstract

Performance within acute medicine services is impacted by ongoing pressures on acute care services. Data from the Society for Acute Medicine Benchmarking Audit 2023 (SAMBA23), was used to assess performance of acute medicine services compared to key clinical quality indicators, comparing performance by initial assessment location. Data was analysed for 8213 unplanned attendances across 161 hospitals. Comparing by initial assessment location, performance against the clinical quality indicators was unchanged from 2022. Only 29% of daytime arrivals assessed within the Emergency Department received consultant review within target times. Delays were seen in transfer between acute care locations. 29% of patients requiring admission were not admitted to the AMU. There is ongoing variation in acute medical service performance nationally, with significant delays in patient access to appropriate assessment locations.

## Introduction

Pathways for patients presenting to acute hospital services remain under pressure, with significant delays frequently experienced by patients within Emergency Departments, including for patients with acute medical problems waiting for admission and assessment by acute medical services.(1–3)

In the UK, assessment and management of patients with an acute medical illness is delivered through acute medical services, accessed via referral from Emergency Medicine or primary and community care services.(4) These processes are delivered within Same Day Emergency Care (SDEC) for patients who can be managed without overnight admission, and Acute Medical Units (AMUs) which can provide rapid investigation and stabilisation for patients at risk of deterioration. Both pathways rely on rapid assessment of patients by clinical decision makers to facilitate appropriate management, whether that is discharge or ongoing care at home or admission to hospital. When last assessed in 2022, performance against clinical quality indicators for acute medicine had deteriorated compared to previous years, with more patients waiting longer than recommended for assessment.(4–7)

We aimed to assess performance of acute medicine services compared to key clinical quality indicators for acute medicine using data from the Society for Acute Medicine Benchmarking Audit 2023 (SAMBA23), and to compare this to previous years. For patients included in SAMBA23, we aimed to assess the interaction between performance against the quality indicators depending on assessment location, and describe delays preceding transfer between locations within emergency and acute medicine services.

## Methods

SAMBA23 took place on Thursday 22^nd^ June 2023. Participation was open to all hospitals accepting unplanned internal medicine admissions (including acute and/or general medicine); community hospitals were excluded. Participating sites registered the audit locally, obtaining appropriate approvals; the SAMBA23 protocol is available online.(8) Data was uploaded to a REDCap database, hosted at the University of Birmingham.(9)

SAMBA23 consisted of a unit level questionnaire, describing hospital and AMU size and unit structure and organisation, and a patient-level questionnaire, completed for all attendances to the medical team over the 24-hour period.

Unplanned medical attendances were assessed against the key clinical quality indicators (CQIs) for acute medicine, recommended by the Society for Acute Medicine (SAM), and based on guidance from the National Institute for Health and Care Excellence (NICE), the Royal College of Physicians (RCP) and Royal College of Physicians of Edinburgh (RCPE).(10–12) These recommend unplanned admissions should have:

- An early warning score recorded within 30 minutes of arrival
- Assessment by a competent clinical decision maker within 4 hours of arrival
- Assessment by consultant physician within either 6 hours if arriving between 8am and 8pm, or 14 hours if arriving between 8pm and 8am.

Location was recorded for the first assessment by a clinical decision maker (whether this was the medical team or another team, including Emergency Medicine (EM)), the first assessment by a clinical decision maker from the medical team, and assessment by consultant physician.

No patient identifiable data is submitted to SAMBA; gender and age (grouped in bands) are recorded. Early warning score was used as a marker of acuity, measured using National Early Warning Score 2 (NEWS2).(13)

For comparison to previous years, 95% confidence intervals for the proportion of patients for whom the target was met were calculated. Comparison between patient group was performed using Chi square tests; a p value of 0.05 was used to signify statistical significance.

## Results

Patient level data was submitted from 166 units at 161 hospitals; multiple registrations at one hospital reflected frailty services registering separately to their main acute medicine service. Of these participating units, 148 submitted unit data alongside patient data. Patient data was submitted from England (141 hospitals), Northern Ireland (6), Scotland (4), Wales (9) and Jersey (1 hospital).

The median number of inpatient beds at participating hospitals was 511 (interquartile range (IQR) 372-718, range 72-1700). The median number of AMU beds per unit was 39 (IQR 28-51, range 10-84). The AMU had a time limit for length of stay in 46% (65/141 hospitals). Thirty-five units (24%) had an enhanced care area/unit within their acute medicine service, containing a median of 8 beds (IQR 5-9, range 3-30).

### Patient level data

Patient level data was submitted for 9612 patients, of which 8213 (85%) were unplanned attendances. The median number of patients seen per unit (planned and unplanned attendances) was 53 (IQR 40-76, range 9-159); the median number of unplanned admissions seen was 46 (IQR 34-63, range 1-119).

### Unplanned attendances

Of the 8213 unplanned attendances, 53% (4314) were female, 47% (3885) were aged 70 years or above and 5.1% (419) were normally resident in a care home (either residential or nursing home). On arrival to hospital, 72% of unplanned admissions had a NEWS2 of 0-2.

The most common source of referral was the Emergency Department (ED, 61%, 4993 patients), followed by general practice (25%, 2036), paramedics (5.5%, 454), other locations within the same hospital (4.9%, 402), via 111 (2.2%,183), and from other hospitals (1.5%,121).

42% of patients (3368/8047) arrived at the hospital via the ambulance service. This varied between units (median 42%, IQR 32-53%, range 0-81%). Patients arriving via the ambulance service were more likely to be aged over 70 (60.1% vs 25.3%, p<0.005), to be male (49.7% vs 45.7%, p=0.001), to be from a care home (10.8% vs 1.1%, p<0.005) and to have a NEWS ≥3 (42.2% vs 17.9%, p<0.005).

Data regarding pre-hospital wait times was recorded for 2559 patients. Of these, 27% had contacted the ambulance service <1 hour before hospital arrival, 45% had contacted them between 1-2 hours before, 23% 2-4 hours before, and 5.7% had first contacted the ambulance service more than 4 hours before hospital arrival.

### Patient location – assessments

65.3% of patients had their initial assessment in the Emergency Department (Table 1). Patients who had their initial assessment in the ED were more likely to have a NEWS2 of 3 or more than those who had their initial clinician assessment in AMU or SDEC (ED: 37%, AMU: 23%, SDEC: 8%, p<0.005).

**Table 1:** Location of key assessment points for unplanned medical admissions. ^1^for some patients, their initial assessment was by the medical team. ^2^3290 patients had their first assessment performed by the medical team ^3^6783 patients required consultant physician review. ED: Emergency Department; AMU: Acute Medical Unit; SDEC: Same Day Emergency Care.

|  | ED |  | AMU |  | SDEC |  | Other locations |  |
| --- | --- | --- | --- | --- | --- | --- | --- | --- |
| Initial assessment<sup>1</sup><br>*Missing: 14* | 5356 | 65.3% | 637 | 7.8% | 2086 | 25.4% | 120 | 1.5% |
| Assessment by medical team<br>*Missing: 45* | 3938 | 48.2% | 1566 | 19.2% | 2426 | 29.7% | 238 | 2.9% |
| Assessment for patients direct to medical team<sup>2</sup><br>*Missing: 5* | 592 | 18.0% | 597 | 18.2% | 2007 | 61.0% | 89 | 2.7% |
| Consultant physician review<sup>3</sup><br>*Missing: 183* | 2226 | 33.7% | 2373 | 36.0% | 1314 | 19.9% | 687 | 10.4% |

First assessment by the medical team occurred in ED in 48.2%, in AMU in 19.2%, and in SDEC in 29.7%. Overall, the percentage of patients who received their medical team assessment in SDEC was higher than in SAMBA22 (24.0%, p<0.005) but varied between units. Twenty units did not assess any unplanned admissions in SDEC. A third or more of admissions were seen by the medical team in SDEC in 35.5% of units (25.4% in SAMBA22).

In the 6612 patients that required consultant review, the most common assessment location was AMU (36.0%)(Table 1).

### Patient location – movement between areas

For patients arriving at or seen within the Emergency Department (data available for 5491 patients), 32.7% were in the ED for less than 6 hours, 35.1% were in the ED for 6-12 hours, and 32.2% were in the ED for more than 12 hours. Seven percent of patients were in the Emergency Department for over 24 hours. Patients who were in the ED for more than 12 hours were more likely to be aged over 70 years (52.2% vs 43.5%, p<0.005), to be a care home resident (7.2% vs 5.7%, p=0.027), and to have a NEWS2 of 3 or more (38.4% vs 32.0%, p<0.005).

For patients seen within SDEC, 1969 patients (80.6%) had data available regarding time before arrival to SDEC; 60% (1182 patients) were transferred to SDEC within 1 hour of hospital arrival. 281 patients were transferred to SDEC but had their first assessment by the ED team: of these, 9.6% (27 patients) were transferred within 1 hour (compared to 68.6% of patients where the medical team delivered their initial assessment, p<0.005) and 43.4% (122 patients) were in the hospital for more than 4 hours before transfer to SDEC (compared to 3.9% of patients where the medical team delivered their initial assessment, p<0.005).

Of the 2442 patients assessed in SDEC, 1439 (58.9%) were not recorded as having any investigations completed prior to transfer. Investigations performed prior to transfer to SDEC are shown in Table 2.

**Table 2:** Investigations completed prior to transfer to SDEC. For all patients seen within SDEC, and for patients referred from ED. SDEC: Same Day Emergency Care; ED: Emergency Department; VBG: venous blood gas; ECG: electrocardiogram.

| Investigation | All patients seen in SDEC<br>(n=2442) |  | Patients seen in SDEC, referred from ED<br>(n=1008) |  |
| --- | --- | --- | --- | --- |
|  | N | % | N | % |
| Point of care blood tests | 141 | 5.8% | 86 | 8.5% |
| Lab blood tests | 775 | 31.7% | 492 | 48.8% |
| VBG | 149 | 6.1% | 110 | 10.9% |
| Chest x-ray | 197 | 8.1% | 139 | 13.8% |
| ECG | 552 | 22.6% | 382 | 37.9% |
| Other imaging | 113 | 4.6% | 59 | 5.9% |
| Rapid Covid-19 swab | 3 | 0.1% | 1 | 0.1% |
| Other | 25 | 1.0% | 12 | 1.2% |

Time waiting in the acute medicine department without a bed was recorded for 418 patients arriving directly to AMU. Of these patients, 28.7% (114 patients) waited more than 4 hours for a bed, and 9.1% (38 patients) waited more than 12 hours.

### Clinical quality indicators

#### Early warning score

73.3% of unplanned admissions had an early warning score recorded within 30 minutes of arrival to hospital (95%CI 72.3-74.2%). Target achievement did not vary comparing patients that had their initial assessment in ED, AMU and SDEC (ED: 74.0%, AMU 73.4%, SDEC: 72.2%, p=0.255). Individual unit performance is shown in Figure 1a. Overall performance against this target was higher than 2022 but lower than previous years (Table 3).

**Figure 1:**
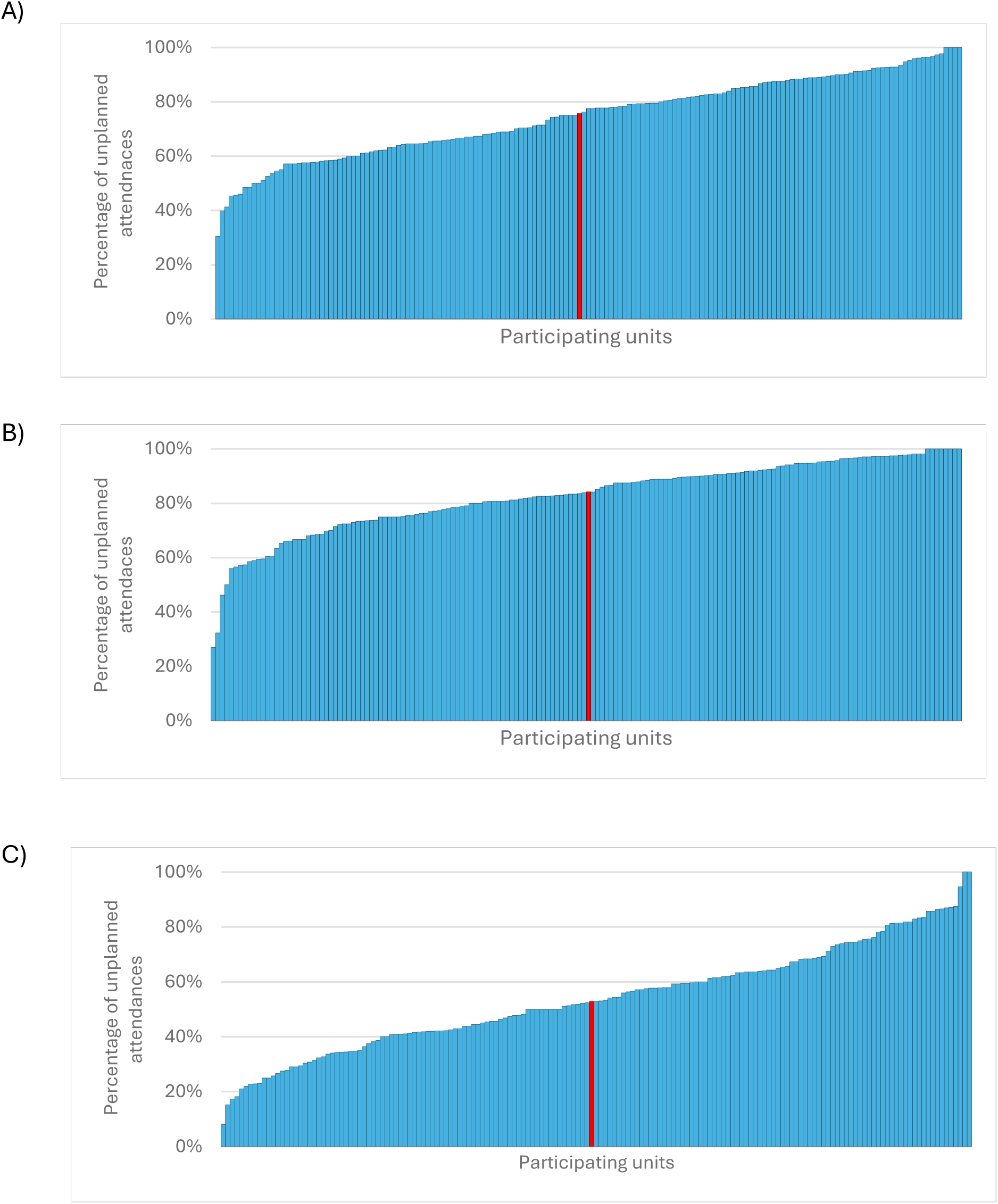
Comparison of unit performance for clinical quality indicators for acute medicine. Units ranked along x-axis, note that units will not be in same order in the three graphs. Median unit performance highlighted in red. A) Percentage of unplanned attendances with early warning score recorded within 30 minutes of arrival; B) Percentage of unplanned attendances assessed by a clinical decision maker within 4 hours of arrival; C) Percentage of patients where consultant review was achieved in the target time (6 hours for arrivals from 08:00-20:00; 14 hours for arrivals from 20:00-08:00).

**Table 3:**
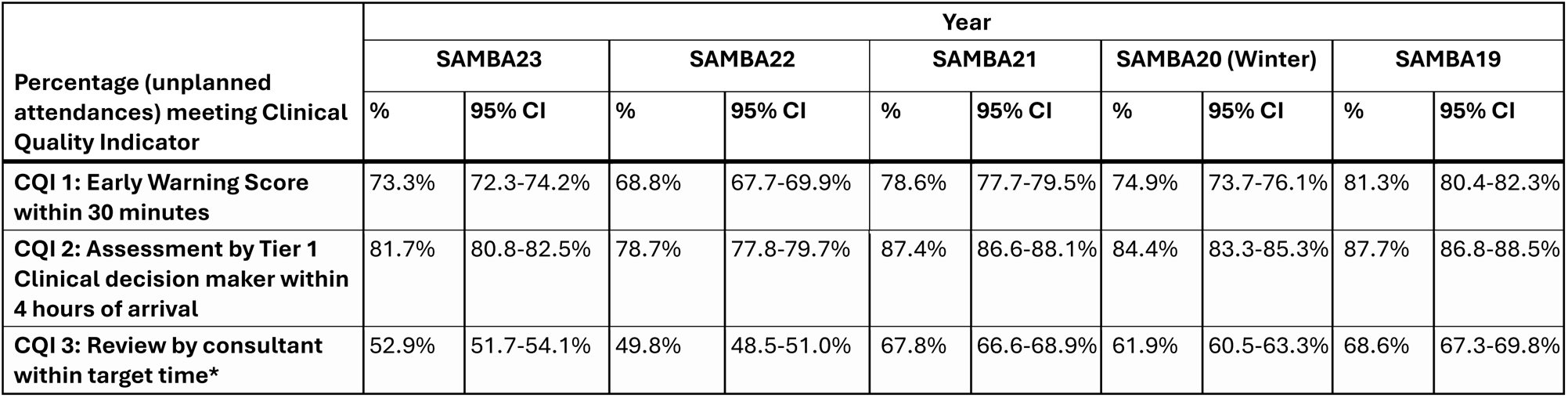
Comparison of performance against Clinical Quality Indicators in SAMBA23 and previous rounds of SAMBA. CQI: Clinical Quality Indicator; CI: confidence interval. *CQI3 target time: 6 hours for patients arriving to hospital from 08:00-19:59; 14 hours for patients arriving to hospital from 20:00-07:59.

| Percentage (unplanned attendances) meeting Clinical Quality Indicator | Year |  |  |  |  |  |  |  |  |  |
| --- | --- | --- | --- | --- | --- | --- | --- | --- | --- | --- |
|  | SAMBA23 |  | SAMBA22 |  | SAMBA21 |  | SAMBA20 (Winter) |  | SAMBA19 |  |
|  | % | 95% CI | % | 95% CI | % | 95% CI | % | 95% CI | % | 95% CI |
| **CQI 1: Early Warning Score within 30 minutes** | 73.3% | 72.3-74.2% | 68.8% | 67.7-69.9% | 78.6% | 77.7-79.5% | 74.9% | 73.7-76.1% | 81.3% | 80.4-82.3% |
| **CQI 2: Assessment by Tier 1 Clinical decision maker within 4 hours of arrival** | 81.7% | 80.8-82.5% | 78.7% | 77.8-79.7% | 87.4% | 86.6-88.1% | 84.4% | 83.3-85.3% | 87.7% | 86.8-88.5% |
| **CQI 3: Review by consultant within target time\*** | 52.9% | 51.7-54.1% | 49.8% | 48.5-51.0% | 67.8% | 66.6-68.9% | 61.9% | 60.5-63.3% | 68.6% | 67.3-69.8% |

#### Assessment by clinical decision maker

81.7% of unplanned admissions were seen by a tier 1 clinician within 4 hours of arrival to hospital (95%CI 80.8-82.5%). Overall performance against this indictor was higher than 2022, but lower than previous years (Table 3). Performance at participating units is shown in Figure 1b. Performance varied with location of assessment, and was more likely to be achieved for patients initially assessed within SDEC services (87.8%) compared to those assessed in ED (79.8%) or AMU (76.7%, p<0.005).

#### Assessment by consultant physician

Overall, 52.9% of unplanned admissions who required a medical consultant review were seen within the target time (95%CI 51.7-54.1%). Performance against this indicator was higher than in 2022, but lower than previous years (Table 3). Comparison of unit performance is shown in Figure 1c. Table 4 shows performance for this indicator by time of arrival and location of initial assessment. The proportion of patients receiving consultant review within the target time was lowest for patients initially assessed in the ED who arrived to hospital between 08:00-20:00. Consultant review was not required for 17% of unplanned admissions.

**Table 4:**
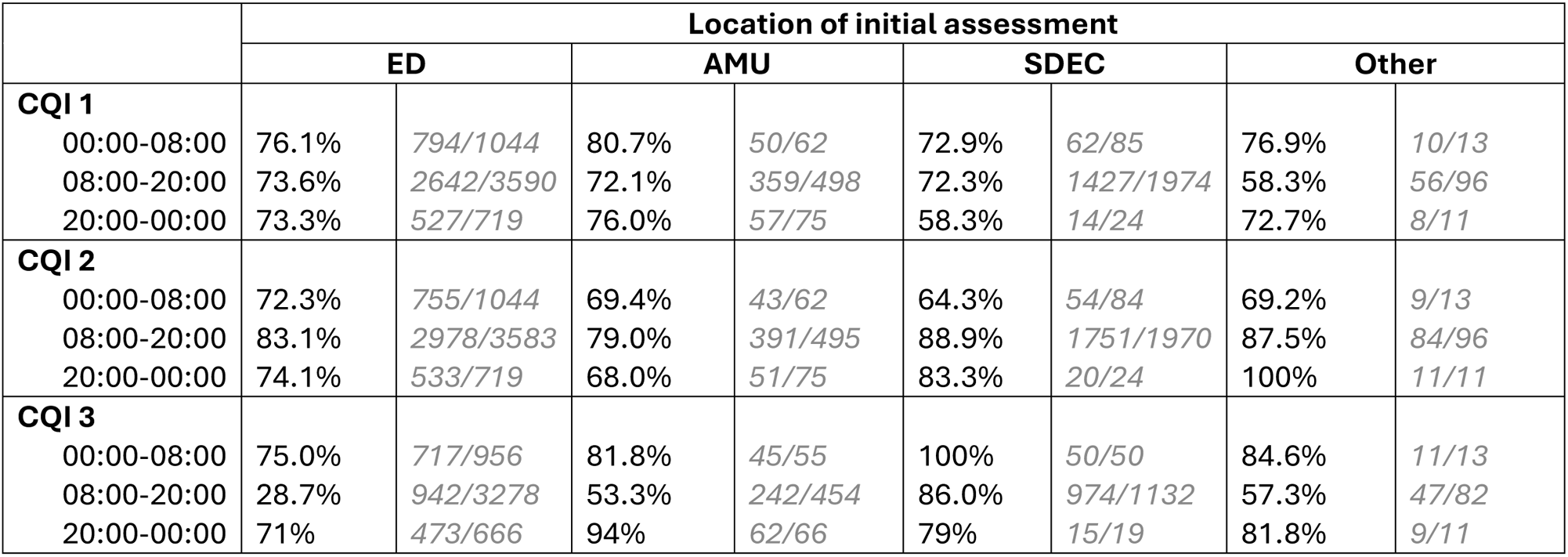
Performance against clinical quality indicators (CQIs) by time of arrival to hospital and initial assessment location. ED: Emergency Department; AMU: Acute Medical Unit; SDEC: Same Day Emergency Care, CQI: Clinical Quality Indicator.

|  | Location of initial assessment |  |  |  |  |  |  |  |
| --- | --- | --- | --- | --- | --- | --- | --- | --- |
|  | ED |  | AMU |  | SDEC |  | Other |  |
| **CQI 1** |  |  |  |  |  |  |  |  |
| 00:00-08:00 | 76.1% | 794/1044 | 80.7% | 50/62 | 72.9% | 62/85 | 76.9% | 10/13 |
| 08:00-20:00 | 73.6% | 2642/3590 | 72.1% | 359/498 | 72.3% | 1427/1974 | 58.3% | 56/96 |
| 20:00-00:00 | 73.3% | 527/719 | 76.0% | 57/75 | 58.3% | 14/24 | 72.7% | 8/11 |
| **CQI 2** |  |  |  |  |  |  |  |  |
| 00:00-08:00 | 72.3% | 755/1044 | 69.4% | 43/62 | 64.3% | 54/84 | 69.2% | 9/13 |
| 08:00-20:00 | 83.1% | 2978/3583 | 79.0% | 391/495 | 88.9% | 1751/1970 | 87.5% | 84/96 |
| 20:00-00:00 | 74.1% | 533/719 | 68.0% | 51/75 | 83.3% | 20/24 | 100% | 11/11 |
| **CQI 3** |  |  |  |  |  |  |  |  |
| 00:00-08:00 | 75.0% | 717/956 | 81.8% | 45/55 | 100% | 50/50 | 84.6% | 11/13 |
| 08:00-20:00 | 28.7% | 942/3278 | 53.3% | 242/454 | 86.0% | 974/1132 | 57.3% | 47/82 |
| 20:00-00:00 | 71% | 473/666 | 94% | 62/66 | 79% | 15/19 | 81.8% | 9/11 |

Comparisons of performance against CQIs by location for daytime arrivals is shown in Table 5. This was unchanged from SAMBA22.

**Table 5:** Performance against clinical quality indicators for daytime arrivals by initial assessment location, comparing SAMBA23 and SAMBA22.

|  | **Year** | **ED** | **AMU** | **SDEC** |
| --- | --- | --- | --- | --- |
| Early Warning Score within 30 minutes | SAMBA23 | 74% | 72% | 72% |
|  | *SAMBA22* | *68%* | *68%* | *73%* |
| Assessment by Tier 1 Clinical decision maker within 4 hours of arrival | SAMBA23 | 83% | 79% | 89% |
|  | *SAMBA22* | *80%* | *80%* | *90%* |
| Review by consultant within target time | SAMBA23 | 29% | 53% | 86% |
|  | *SAMBA22* | *28%* | *53%* | *87%* |

### Outcomes after 7 days

32.9% of unplanned medical attendances were discharged on the same day, 39.0% were discharged within the next 7 days, 22.4% remained in hospital at more than seven days. The mortality within 7 days was 2.2%.

Of the patients assessed within SDEC services, 82.6% of patients were discharged same day. The proportion of patients assessed within SDEC and discharged the same day varied between services (median 85.2%, IQR 75-97.7%, range 0-100%). Of those discharged on the day of arrival, 72.7% had their first medical team assessment in SDEC services.

Overall, 41.9% of patients (2969/7093 with data available) did not spend any time on AMU during their attendance; excluding patients assessed within SDEC, 29.2% of patients (1535/5264) did not spend any time on AMU. Of 4124 patients where time on AMU was reported, 514 (12.5%) spent more than 72 hours on the AMU.

## Discussion

These results suggest that although overall performance against key clinical quality indicators for acute medical services has improved from 2022, performance remains lower than previous years. However, comparison by initial assessment location suggests performance is unchanged in comparison to 2022, with apparent improvements driven by an increase in the proportion of patients assessed within Same Day Emergency Care services.

Delivery of care within the time frames outlined by the clinical quality indicators is assumed to represent patient access to appropriate assessment, investigation and treatment in a timely manner. Therefore, continued reduced performance against these indicators reflects ongoing delays in necessary care for a considerable proportion of medical patients. In addition, these results demonstrate that many patients are experiencing delays in transfer to appropriate assessment locations, spending prolonged periods within the Emergency Department before transfer to acute medicine services, whether for admission to an AMU or for assessment within SDEC services.

Although unplanned medical admissions are the most common cause of hospital admission, delays within their admission pathways are not routinely monitored within national data, beyond assessment of targets relating to the delivery of care within emergency departments.(14) In this analysis, a third of medical patients spent more than 12 hours within the Emergency Department. These patients were likely to be older, and at higher risk of deterioration, as assessed by NEWS2.(13) Previous analysis from the Royal College of Emergency Medicine found that almost 20% of patients were in the ED for more than 6 hours from hospital arrival, higher than nationally reported figures measured from decision to admit rather than from arrival;(15) medical patients may be more likely than the overall ED cohort to experience prolonged waits, due to delays awaiting inpatient bed availability. These delays have multiple impacts, preventing patients from accessing the benefits associated with care delivered on AMUs, as well as increasing the nursing burden on ED staff, and contributing to ED overcrowding, with knock on impacts to emergency care delivery. Most importantly, these delays have been shown to be associated with increased mortality.(16) Collaboration between Emergency Medicine, acute medicine and other hospital internal medicine services should be encouraged and supported in order to address these issues.

Performance against clinical quality indicators was measured from arrival to hospital, as in previous rounds of data collection, to reflect patient experience. Although the recommended quality indicators suggest performance can be measured from patient arrival to AMU, after excluding patients assessed through SDEC, almost 30% of patients assessed by the medical team were not transferred to the AMU on this admission. This limits the utility of arrival to AMU in assessing acute medical service performance currently, but more importantly it suggests that a substantial proportion of patients with medical emergencies do not have access to the benefits provided by AMU care.(17)

Although the proportion of patients assessed through SDEC services has increased from previously, there remains considerable variation between hospitals in the proportion of patients assessed within SDEC services. The NHS Long Term Plan suggests that one third of patients be managed without admission to an inpatient bed; to deliver this target through SDEC services, at least one third of attendances would need to be assessed within SDEC, a target met by only 35% of participating hospitals.(18) A third of unplanned attendances were discharged without overnight admission, but a quarter of these patients were not assessed within SDEC services, providing further evidence that a portion of patients are not reaching the most suitable location for their needs.

This analysis has some limitations. Participation in SAMBA is voluntary, and not all acute medicine services participate in SAMBA; there may be systematic differences in the units that participate and those that do not. There is a higher response rate in England compared to the other UK nations. The patient-level data represents a single day of care, and there may be variation in performance over time. Longitudinal data analysis may be beneficial, but is not currently reported within national datasets.

For assessment of time intervals spent within locations in the acute care pathway, there were varying proportions of missing data, which should be considered when interpreting these results. This missing data may reflect variation in ease of access to this data, for example due to availability of electronic data or access across systems. This highlights the difficulties that acute services may encounter when trying to assess the patient journey and experience as a whole. The analysis presented here highlights the delays that some patients are experiencing at all stages of the admission process, including in the pre-hospital interval while awaiting admission via ambulance services, and within the hospital setting while awaiting transfer to the most appropriate admission service and to assessment.

The high proportion of consultant physician reviews in other locations likely reflects movement to inpatient wards. Other acute medicine assessment locations, where participating units felt that their acute medicine location was not an AMU or SDEC area, were included in ‘other locations’; further exploration of the perceived definitions of AMU and SDEC services may help to improve understanding of the range of service structure used within acute services.

## Conclusion

Performance against clinical quality indicators within acute medicine services remains lower than 2019-2021. There remains considerable variation in performance, and significant delays in patient access to the assessment locations most likely to be beneficial to the management of their acute medical illness. Almost a third of patients who are admitted do not benefit from the rapid care coordination provided by the Acute Medical Unit.

## Data Availability

All data produced in the present study are available upon reasonable request to the authors

## References

1. The King’s Fund. What’s going on with A&E waiting times? [Internet]. 2022 [cited 2024 May 28]. Available from: https://www.kingsfund.org.uk/insight-and-analysis/long-reads/whats-going-on-with-ae-waiting-times

2. Royal College of Emergency Medicine. Data show 1.65 million patients in England faced 12-hour waits from time of arrival in A&Es in 2022. 2023 Feb 28 [cited 2024 May 28]; Available from: https://rcem.ac.uk/data-show-1-65-million-patients-in-england-faced-12-hour-waits-from-time-of-arrival-in-aes-in-2022/

3. Cooksley T, Holland M, Sapey E. Reversing the urgent and emergency care spiral of decline. BMJ. 2023 Jul 3;p1530.

4. Atkin C, Knight T, Cooksley T, Holland M, Subbe C, Kennedy A, et al. Performance of admission pathways within acute medicine services: Analysis from the Society for Acute Medicine Benchmarking Audit 2022 and comparison with performance 2019 - 2021. Eur J Intern Med. 2023 Dec;118:89–97.

5. Holland M, Subbe C, Atkin C, Knight T, Cooksley T, Lasserson D. Society for Acute Medicine Benchmarking Audit 2019 (SAMBA19): Trends in Acute Medical Care. Acute Med. 2020;19(4):209–19.

6. Atkin C, Knight T, Subbe C, Holland M, Cooksley T, Lasserson D. Acute care service performance during winter: report from the winter SAMBA 2020 national audit of acute care. Acute Med. 2020;19(4):220–9.

7. Atkin C, Knight T, Cooksley T, Holland M, Subbe C, Kennedy A, et al. Society for Acute Medicine Benchmarking Audit 2021 (SAMBA21): assessing national performance of acute medicine services. Acute Med J. 2022 Jan 1;21(1):19–26.

8. Society for Acute Medicine. SAMBA [Internet]. 2024. Available from: https://www.acutemedicine.org.uk/samba-new/

9. Harris PA, Taylor R, Thielke R, Payne J, Gonzalez N, Conde JG. Research electronic data capture (REDCap)—A metadata-driven methodology and workflow process for providing translational research informatics support. J Biomed Inform. 2009 Apr;42(2):377–81.

10. Society for Acute Medicine. Clinical Quality Indicators for Acute Medical Units. [Internet]. 2011 [cited 2024 May 28]. Available from: https://www.acutemedicine.org.uk/blog/2010/11/15/acute-care-quality-and-performance-indicators/

11. Langlands A, Dowdle R, Elliott A, Gaddie J, Graham A, Johnson G, et al. RCPE UK Consensus Statement on Acute Medicine, November 2008. Br J Hosp Med Lond Engl 2005. 2009 Jan;70(1 Suppl 1):S6–7.

12. National Institute for Health and Care Excellence. NICE Guideline NG94: Emergency and acute medical care in over 16s: service delivery and organisation. 2018.

13. Royal College of Physicians. National Early Warning Score (NEWS) 2 [Internet]. 2017. Available from: https://www.rcplondon.ac.uk/projects/outputs/national-early-warning-score-news-2

14. NHS England. Hospital Accident & Emergency Activity [Internet]. [cited 2024 May 28]. Available from: https://digital.nhs.uk/data-and-information/publications/statistical/hospital-accident--emergency-activity

15. Royal College of Emergency Medicine. Tip of the Iceberg: 12 hour stays in the Emergency Department [Internet]. 2022 [cited 2024 May 28]. Available from: https://rcem.ac.uk/wp-content/uploads/2022/06/Tip-of-the-Iceberg-12-Hour-Stays-in-the-Emergency-Department.pdf

16. Jones S, Moulton C, Swift S, Molyneux P, Black S, Mason N, et al. Association between delays to patient admission from the emergency department and all-cause 30-day mortality. Emerg Med J. 2022 Mar 1;39(3):168.

17. Scott I, Vaughan L, Bell D. Effectiveness of acute medical units in hospitals: a systematic review. Int J Qual Health Care. 2009 Dec 1;21(6):397–407.

18. National Health Service. The NHS Long Term Plan [Internet]. 2019. Available from: https://www.longtermplan.nhs.uk/wp-content/uploads/2019/08/nhs-long-term-plan-version-1.2.pdf

